# Influenza-Negative Influenza-Like Illness (fnILI) Z-Score as a Proxy for Incidence and Mortality of COVID-19

**DOI:** 10.1101/2020.04.22.20075770

**Authors:** Fatima N. Mirza, Amyn A. Malik, Saad B. Omer

## Abstract

Though ideal for determining the burden of disease, SARS-CoV2 test shortages preclude its implementation as a robust surveillance system in the US. We correlated the use of the derivative influenza-negative influenza-like illness (fnILI) z-score from the CDC as a proxy for incident cases and disease-specific deaths. For every unit increase of fnILI z-score, the number of cases increased by 70.2 (95%CI[5.1,135.3]) and number of deaths increased by 2.1 (95%CI[1.0,3.2]). FnILI data may serve as an accurate outcome measurement to track the spread of the and allow for informed and timely decision-making on public health interventions.

## Background

Severe acute respiratory syndrome coronavirus 2 (SARS-CoV-2) has spread exponentially since December 2019, transforming from a localized outbreak in Wuhan, China to a global pandemic. As of mid-April, roughly 2 million cases have been reported(1), with one-fifth of cases requiring inpatient hospitalization and a case-fatality at an estimated 2%(2).

As the number of individuals infected rapidly climbs, the testing capacity has been outpaced by the need for such tests. In the United States, this has posed a challenge to physicians and public health professionals at large, particularly as it relates to accurately tracking the spread of disease. Assessing the intensity of the epidemic nationally in a given region is the backbone of allocating resources at the federal and state level, and inform the implementation or relaxing of public health restrictions (e.g. initiating or easing a lockdown).

Given the rapid increase in cases in the previous weeks without parallel expansion in testing capacity and unclear specificity/sensitivity, this problem will only continue to be exacerbated until a nationwide program is made available and further validation studies have been completed (3). In the interim, there is an urgent need to identify proxies for disease incidence that are routinely collected through available infrastructure in the United States in order to guide the evolving public health response in this country(4).

The Centers for Disease Control and Prevention (CDC) centrally collates data using the U.S. Outpatient Influenza-like Illness Surveillance Network (ILINet) and the National Respiratory and Enteric Virus Surveillance System (NREVSS) (5). We believe that combining both sources of this publicly-available, routinely collected data may serve as a reliable proxy for SARS-CoV-2 incidence and mortality. In this study, we used influenza-negative ILI (fnILI) z-scores and compared them against the reported COVID-19 cases and deaths by week to document trends over time.

## Methods

We downloaded flu negative influenza-like illness (fnILI) data derived from the Center for Disease Control and Prevention’s ILINet and NREVSS data for states (5,6). ILINet consists of records from outpatient healthcare providers in all 50 states and reported 60 million patient visits in the 2018-2019 season. Weekly, approximately 2,600 outpatient healthcare providers around the country report the total number of patients as well as those with influenza-like illness, defined as a temperature of 100^°^F or greater alongside a cough and/or sore throat, as well as regional baseline are reported. These data are weighted by state population, and percentage of flu-positive influenza-like illness is compared to regional estimates and a historical nationwide baseline of 2.4%. NREVSS provides virologic surveillance data weekly from approximately 100 public health and over 300 clinical laboratories throughout the United States, including the total number of respiratory specimens tested for influenza, the number positive for influenza viruses, and the percent positive by influenza virus type. Some states may have limited data or have delays in reporting that may not make this information immediately available.

Reich et al (6) reviewed twenty-three seasons of influenza data, beginning in 1997, and ten seasons of statewide data, beginning in 2010 and calculated fnILI from the CDC (5). fnILI was determined using weighted influenza-like illness (wILI) from ILINet – which represents the percentage of doctor’s office visits that presented with a primary complaint of fever and one additional influenza-like symptoms – and percentages of positive influenza specimens from NREVSS data, compared to a baseline calculated from prior all seasons of data extracted as described above. fnILI was calculated as:

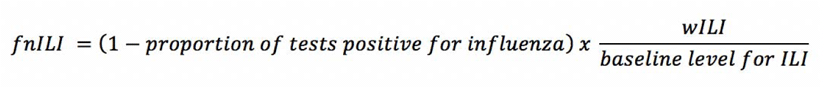

These data included a z-score that represents the degree to which a given fnILI observation was significantly lower or higher than expected based on past trends at similar times during prior years. Z-score was calculated as:

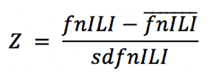

with as the average weekly observations for the past nine years with one week on either side and as associated standard deviation.

We merged this dataset with the CDC-reported SARS-CoV-2 cases and disease-specific deaths. We graphically represented the fnILI z-score, cumulative cases, and cumulative deaths for the contiguous U.S for the month of March 2020. We used a mixed effects linear model accounting for clustering at the state level using random effects to determine the relationship between weekly case counts or deaths and fnILI z-score, and a lag term to account for delay between onset of symptoms and confirmation of diagnosis. We selected the model with the lowest Akaike Information Criteria (AIC) and Bayesian Information Criteria (BIC) (7) while maintaining statistical significance. Median fnILI z-score across all states were determined by week and plotted against total nationwide cases and/or deaths per week. All data was analyzed using Stata v15.0 (StataCorp, College Station, Texas).

## Results

fnILI data was available for all states except Florida, New Hampshire, New Jersey, and Rhode Island. Over the course of the study period, September 29th 2019 to March 29th 2020, there were 123,509 reported COVID-19 cases and 2,303 deaths, representing a case-fatality rate of 1.8%. This represented 36,749,916 unique patient visits across the US with an average of 3,040 providers nationwide per week. The rate of fnILI across the study period was 85.8% and the average flu positive baseline across the country was 1.4%.

There is an apparent tracking between fnILI z-score and cases or deaths by state over the course of the study period. This phenomenon is particularly pronounced when comparing these indices during the month of March (Figure 1A).

**Figure 1.**
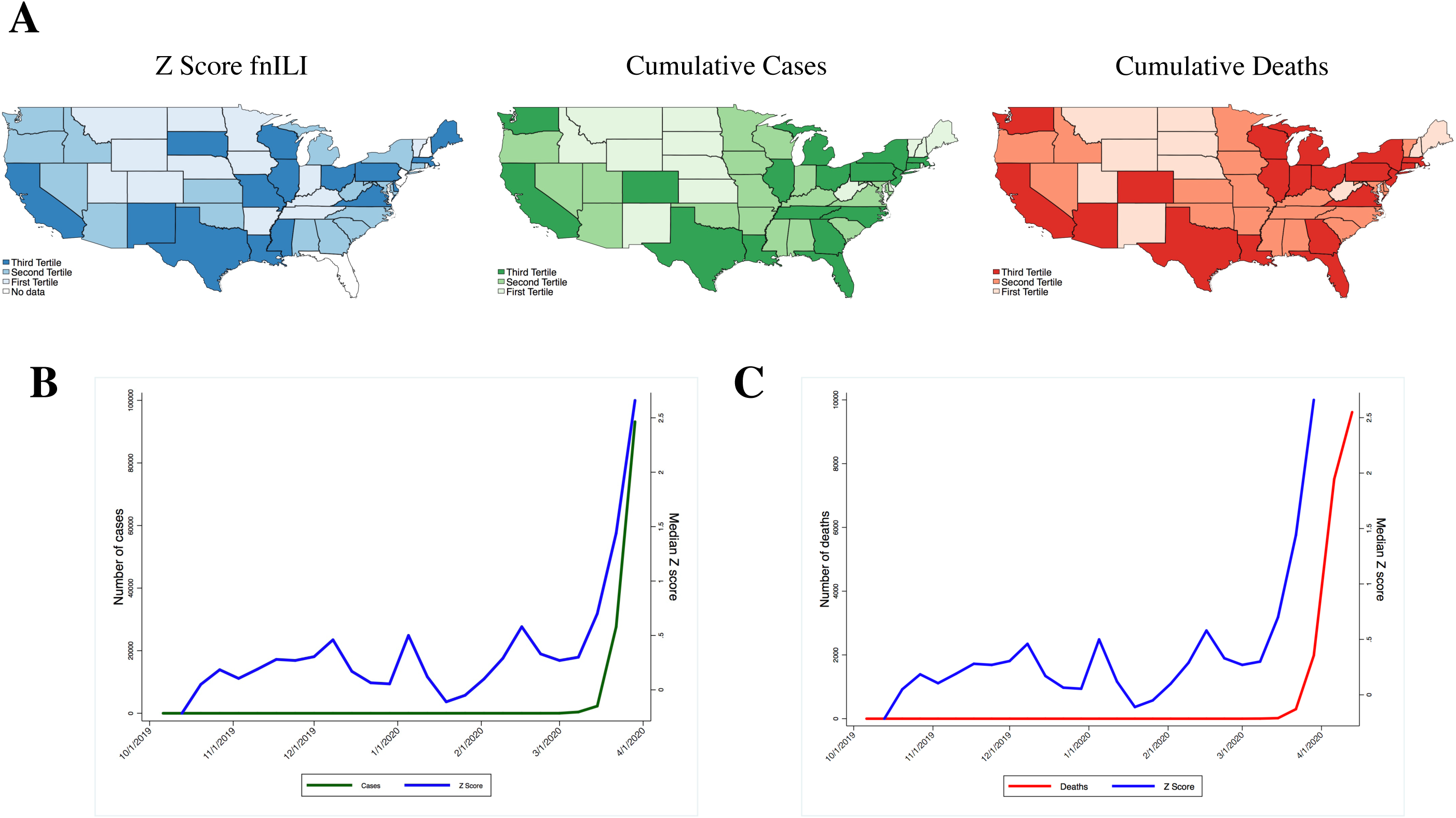
Overall state-specific and median nationwide fnILI Z-Score versus COVID-19 cases or deaths. (A) fnILI z-scores and cumulative cases/deaths for individual states in the month of March. (B) Nationwide weekly median fnILI z-scores versus new cases nationwide. (C) Nationwide weekly median fnILI z-scores versus deaths nationwide.

When assessing the correlation over time between fnILI z-score and either new cases or deaths, we observed a z-score peak prior to an increase in cases or deaths. Therefore, we used a lag variable of two weeks for incidence and one week for mortality to better fit the model (Table 1). On the mixed effects linear model accounting for clustering at the state level using random effects, we found that for every unit increase of fnILI z-score two weeks prior, the number of cases increased by 70.2 (95% CI [5.1, 135.3]). Similarly, we found that for every unit increase of fnILI z-score one week prior, the number of deaths increased by 2.1 (95% CI [1.0, 3.2]), also when correcting for regional effects. When plotting the median nation-wide z-score two week or one week prior, respectively, versus new cases (Figure 1B) or deaths (Figure 1C), the two measures tracked. Nationwide, one unit increase of fnILI z-score correlated with 49243.4 (95% CI[34250.8, 64235.9]) and 608.3 deaths (95% CI[477.3, 739.4]).

**Table 1.** Possible Models for fnILI Z-Score and Incidence or Mortality.

| Incidence | Lag | Coef. | 95% CI |  | p | AIC | BIC |
| --- | --- | --- | --- | --- | --- | --- | --- |
|  |  |  | Lower | Upper |  |  |  |
|  | None | 128.9 | 79.2 | 178.6 | <0.01 | 20516.2 | 20536.5 |
|  | 1 Week | 129.9 | 75.5 | 184.3 | <0.01 | 19834.7 | 19854.8 |
|  | 2 Week | 70.2 | 5.1 | 135.3 | 0.03 | 19095.9 | 19115.9 |
|  | 3 Week | 1.97 | -68.4 | 72.3 | 0.9 | 18364.2 | 18384 |
| Mortality | None | 2.1 | 1.1 | 3.1 | <0.01 | 11328.3 | 11348.6 |
|  | 1 Week | 2.1 | 1.0 | 3.2 | <0.01 | 10970.8 | 10991.0 |
|  | 2 Week | 1.2 | -0.08 | 2.5 | 0.06 | 10585.1 | 10605.1 |

## Discussion

There is increasing evidence that current testing may significantly underestimate the burden of disease (8), necessitating alternative methods to accurately assess these trends over time. Our results suggest that the fnILI z-score data can be used as a proxy for the trajectory of disease incidence and mortality in the United States. In the context of limited resources in a rapidly changing field, it becomes increasingly necessary to innovatively utilize available infrastructure to tackle the apparent gaps in knowledge quickly. To our knowledge, this is the first academic study to use fnILI z-scores from ILINet and NVRESS data in order to model and potentially predict the burden of COVID-19 over time.

This report demonstrates the important potential of such a proxy, and validates its correlation with incidence and mortality. Importantly, we present the optimal model for such a prediction by building in a lag term. This two week lag term is likely necessary for incidence due to the known incubation period of this disease and because of a delay in testing (2). For mortality, our one week lag term likely represents the rapid escalation to fatality (9). These lag terms also may allow our model to function as an early warning system for rise in cases, similar to ILINet.

As there is already a robust infrastructure in place to collect these data, validating the use of such data is extremely valuable, especially in the setting of limited availability and capacity of testing kits. fnILI provides a good proxy in the absence of testing to evaluate the results of public health interventions and make timely decisions to change course.

## Data Availability

All data is available on the Center for Disease Control and Prevention’s website and by Reich et al.

https://www.cdc.gov/flu/weekly/overview.htm

https://github.com/reichlab/ncov/blob/master/analyses/ili-labtest-report-20200403.pdf

## Footnote

### Disclosures

The authors have no conflicts of interests to disclose.

### Funding/Support

There is no funding for this study.

### Statement of Prior Presentation

This information has not been previously presented.

